# Almond Consumption Improves Inflammatory Profiles Independent of Weight Change: A 6-Week Randomized Controlled Trial in Adults with Obesity

**DOI:** 10.1101/2025.09.08.25334937

**Authors:** Ayodeji Adepoju, Elaheh Rabbani, Philip Brickey, Victoria Vieira-Potter, Jaapna Dhillon

## Abstract

**Background:** Obesity is characterized by chronic low-grade systemic inflammation that contributes to metabolic dysfunction. Diet is a modifiable factor that can help reduce this inflammation. Nuts such as almonds are rich in unsaturated fats, and antioxidant and anti-inflammatory micronutrients, which may work synergistically to attenuate obesity-related inflammation. Hence, the objective of this study was to investigate whether daily almond consumption improves systemic inflammatory and immune markers in adults with obesity.

**Methods:** In this randomized controlled parallel-arm trial (ClinicalTrials.gov ID: NCT05530499), 69 adults (age: 30-45 years) with obesity (BMI: 30-45 kg/m^2^) were assigned to consume either 57 g/day of almonds (n = 38) or an isocaloric snack (cookie; n = 31) for six weeks. Fasting serum inflammatory cytokines, innate immune cell counts, body weight, serum glucose, insulin, lipid profile, and alpha-tocopherol were measured at baseline and week six. Dietary intake, compliance, palatability, acceptance, and appetite ratings were also assessed. Primary outcomes were analyzed using linear mixed models and baseline-adjusted linear models.

**Results:** Compliance was high in both groups, with greater acceptance of almonds (P<0.05); however, serum alpha-tocopherol did not change. Almond consumption significantly decreased serum IL-6, TNF-α, and IFN-γ over 6 weeks compared with the cookie group (P<0.05). No significant group differences were observed for innate immune cell counts, body weight, appetite ratings, blood pressure, or serum fasting glucose, insulin, total cholesterol (C), LDL-C, and triglycerides over six weeks. The almond group also increased intakes of monounsaturated fat, fiber, alpha-tocopherol, magnesium, zinc, and manganese, and improved diet quality indices (P<0.05).

**Conclusion:** Daily almond consumption for six weeks improved inflammatory cytokine profiles in adults with obesity, without changes in body weight under free-living conditions. These findings support recommending almonds as part of healthy dietary patterns to help attenuate obesity-related inflammation.

## 1. Introduction

Obesity is a state of chronic low-grade systemic inflammation, characterized by elevated circulating concentrations of pro-inflammatory cytokines and measurable shifts in innate immune cell populations [1,2]. Adipose tissue expansion in obesity amplifies inflammatory signaling contributing to immune cell activation which eventually exacerbates systemic inflammation [1–3]. This persistent inflammatory response is a key biological mechanism linking obesity to increased risk of insulin resistance, type 2 diabetes, and cardiovascular complications [4–7].

Diet is a modifiable factor that can help reduce the chronic low-grade systemic inflammation observed in obesity. Among various healthy eating patterns, the Mediterranean (MED)-style dietary pattern has consistently demonstrated beneficial effects on inflammatory and immune markers. Large epidemiological studies, including the Nurses’ Health Study and the ATTICA study, report that higher adherence to a MED-style dietary pattern is associated with lower concentrations of pro-inflammatory cytokines and immune cell counts [8,9]. For example, findings from the ATTICA study showed that participants with the highest adherence had on average, 17% lower IL-6 concentrations and 14% lower white blood cell counts compared to those with the lowest adherence. Systematic reviews and meta-analyses of randomized controlled trials (RCTs) further support that MED-style dietary interventions reduce circulating inflammatory markers such as IL-6, IL-1β, and C-reactive protein (CRP) across diverse populations [10,11]. Another systematic review on plant-based diets shows a reduction of IL-6 but not TNF-alpha for individuals with overweight and obesity with diverse health status such as metabolic syndrome, CVD, or diabetes [12].

Nuts are a core component of high-quality dietary patterns [13] and may contribute substantially to the anti-inflammatory effects associated with these dietary patterns. Almonds, in particular, are among the most widely consumed tree nuts globally [14] and provide a variety of nutrients and bioactive compounds, including monounsaturated fats, vitamin E, zinc, magnesium, polyphenols, that are important for regulating inflammation, immune function, and oxidative stress [15–17]. To date, most evidence on almonds and inflammation has been synthesized from studies in populations with diverse BMI and health status [18–20], yielding mixed results and limiting obesity-stratified inferences for inflammatory markers. Addressing these gaps could strengthen dietary recommendations aimed at lowering obesity-related inflammation and its metabolic consequences. Therefore, the primary objective of this randomized, controlled, parallel-arm clinical trial was to investigate the effects of six weeks of almond consumption on systemic immune and inflammatory markers in adults with obesity.

## 2. Materials and Methods

The procedures involving human participants in this study were approved by the Institutional Review Board of the University of Missouri-Columbia (Protocol number: 2090290). The study is registered on ClinicalTrials.gov (NCT05530499).

### 2.1 Participant Characteristics

Participants were recruited via public advertisements. Inclusion criteria included the following: (a) age 30–45 years old, (b) BMI of 30 – 45 kg/m^2^, (c) willingness to maintain consistent diet and activity patterns, (d) willingness to consume study foods, (e) non-smokers over the past year, (e) and weight stable (no greater than 5 kg change over the last 3 months). Exclusion criteria included diagnosed diabetes (fasting blood glucose >125 mg/dL), pregnant or lactating individuals, regular consumer of nuts (>3 times/week), planning on getting viral vaccines during study, allergies to study foods, illicit drug use, current use of medication that interfere with immune measures, recent start of medications that affect metabolism or appetite and drug therapy for coronary artery disease, peripheral artery disease, congestive heart failure or dyslipidemia. A total of 90 individuals were screened to determine eligibility for participation in this study, and 69 participants started the study. The participant flow is depicted in **Figure 1**.

**Figure 1:**
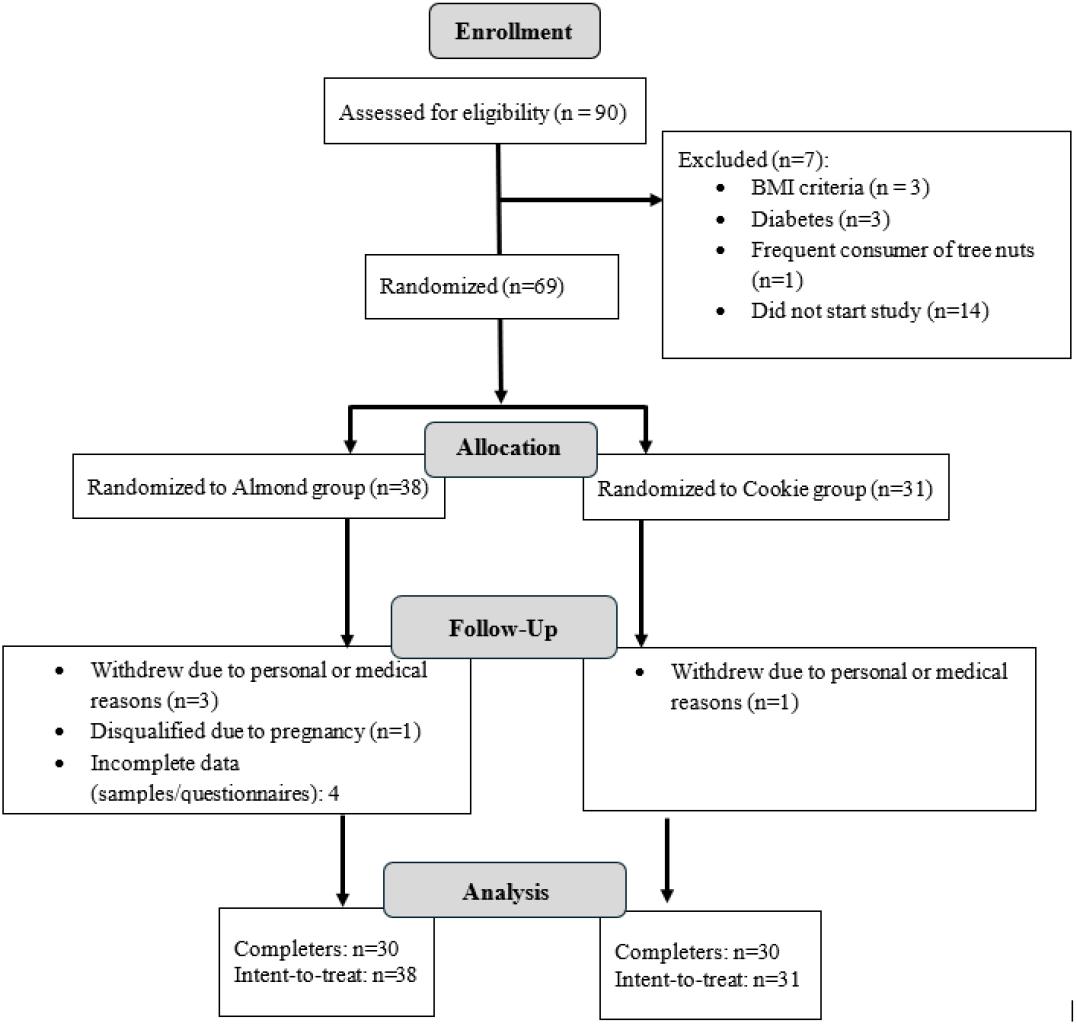
Consort flow diagram of participants in the almond and cookie groups from enrollment to study completion.

### 2.2 Study Design

The study was a 6-week randomized, controlled, parallel-arm intervention. Participants were randomly assigned into one of two study arms: whole natural dry-roasted almonds (n = 38; 57 g/day (2 oz); 322 kcal) or isocaloric comparator of cookies (n=31; 66g (6 cookies); 325 kcal). The cookie was selected as the comparator because it represents a typical processed snack consumed in North America and was intended to reflect existing dietary exposure in this target population [21–23]. The nutrient composition of the study foods is shown in **Table S1**. Participants incorporated the assigned foods into their usual diet according to personal preference and maintained their customary dietary and physical activity habits. No dietary counseling was provided in this free-living trial. Study foods were dispensed at baseline and again at the week-3 check-in visit. Compliance with study food consumption was monitored via daily text messages asking participants whether they consumed their foods and the time of consumption (morning, afternoon, evening). The baseline characteristics of the participants are shown in **Table 1**.

**Table 1.** Baseline characteristics of participants in the almond and cookie groups in the 6-week randomized controlled trial in adults with obesity.

| Characteristic | Almond | Cookie |
| --- | --- | --- |
| Sample size, n | 38 | 31 |
| Sex, n (%) |  |  |
| Female | 31 (82%) | 20 (65%) |
| Male | 7 (18%) | 11 (35%) |
| Age<sup>§</sup>, years | 37.8 ± 4.3 | 36.4 ± 4.4 |
| BMI<sup>§</sup>, kg/m² | 35.9 ± 4.3 | 34.8 ± 3.4 |
| Race, n (%) |  |  |
| White | 30 (79%) | 27 (87%) |
| Black or African American | 8 (21%) | 2 (6.5%) |
| Asian | - | 2 (6.5%) |
| Ethnicity, n (%) |  |  |
| Hispanic or Latino | 2 (5%) | 3 (10%) |
| Not Hispanic or Latino | 36 (95%) | 28 (90%) |

§, Mean ± SD

### 2.3 Sample Size Calculation and Group Randomizations

Sample size calculations for this study were informed by preliminary data from a previous intervention [24] involving a subset (n=20) of participants per group (almond vs. cracker) in young adults with mixed BMI status. In that population, we observed Cohen’s d effect sizes ranging from small to moderate (d=0.19-0.60) for 8-wk changes in key markers such as IL-6, IL-10, and TNF-alpha. Given that the current study targets a population of middle-aged adults with obesity, who typically exhibit greater baseline inflammation [25–29], we anticipated at least equal or larger effect sizes than those observed in the earlier cohort. Assuming a moderate effect size (Cohen’s d = 0.6), a two-tailed alpha of 0.05, and 80% power, a sample size of approximately 25-30 participants per group was estimated to be sufficient to detect differences over 6 weeks in most markers. To account for anticipated attrition (∼10–20%), we aimed to enroll approximately 34–38 participants per group to achieve ∼30 completers per group. Randomization to the groups was performed using a restricted 1:1 allocation sequence generated in Microsoft Excel. A list containing 42 almond and 42 cookie assignments was randomly permuted using the RAND() function, and participants were assigned sequentially. Enrollment was stopped once 30 participants per group completed the study. This was a single-blinded trial, where clinical and laboratory analysts were blinded to group allocation. Blinding was achieved by coding samples with ID numbers.

### 2.4 Outcomes

Primary outcomes for this study were immune and inflammatory markers. Secondary outcomes for this analysis were anthropometric, cardiovascular, dietary, and appetite ratings. All outcomes were assessed before and after the 6-week study at minimum.

#### 2.4.1. Blood Immune and Inflammatory Markers

Blood samples were obtained at each visit using butterfly needle venipuncture after at least 8 hours of fasting. Blood samples for white blood cells (WBC) assessments were collected in EDTA tubes. Differential WBC count i.e., absolute and proportions for lymphocytes, neutrophils, monocytes, eosinophils, and basophils were assessed using counting and microscopy methodologies by Quest Diagnostics-Lenexa in Kansas.

Serum samples were obtained from blood collected in serum separator tubes (SST) which were allowed to sit for 1 hour before centrifugation for 10 min at 1.3 RCF. Serum samples were stored at −80 °C until analysis. Fasting serum concentrations of inflammatory markers TNF-alpha (Kit no.: KAC1751), IFN-gamma (KAC1231), IL-6 (KAC1261), and IL-10 (KAC1321) were quantified using commercially available ELISA kits from Invitrogen.

#### 2.4.2 Blood Metabolic and Other Markers

Fasting serum glucose concentrations were measured using an enzymatic amperometric glucose oxidase method by the YSI 2300 STAT Plus device (Yellow Springs, OH). Fasting serum triglycerides (TG; E-BC-K238), total cholesterol (TC; E-BC-K109-S), and high-density lipoprotein-cholesterol (HDL-C; E-BC-K221-M) concentrations were measured using commercially available enzymatic colorimetric assays (Elabscience^®^, USA). Low-density lipoprotein-cholesterol (LDL-C) was calculated using Friedwald equation LDL-C = [(TC – HDL-C) – (TG/2.2)] [30]. The fasting serum concentrations of insulin was quantified using commercially available ELISA kit from Millipore (EZHI-14k). Insulin sensitivity and resistance indices were calculated as we have done previously [31]. Quantitative insulin-sensitivity check index (QUICKI), an estimate of fasting insulin sensitivity, was computed using [1/[log(fasting insulin μU/mL) + log(fasting glucose mg/dL)]. Homeostatic model assessment for insulin resistance (HOMA-IR), an estimate of fasting insulin resistance was computed using [fasting insulin (µU/L) × fasting glucose (mg/dL)/405].

Blood samples for tocopherol measurements were collected in light-protected tubes and transported to a Quest Diagnostics^®^ laboratory for analysis at the end of the participant visit. Tocopherol concentrations were assessed using chromatography methodology by SLI Quest Diagnostics–Nichols Valencia in California.

#### 2.4.3. Anthropometric and Blood Pressure Outcomes

Height was measured to the nearest 1 mm using a stadiometer (SECA 216 Height Measuring Rod, SECA). Body weight was assessed in lightweight clothes to the nearest 100 g using standard calibrated electronic scales. Waist and hip circumferences were recorded at the narrowest part of the torso and the widest part of the hips-gluteal region respectively using a standard measuring tape. Resting systolic and diastolic blood pressure (BP) was assessed using an OMRON automatic BP device (model BP:725, Kyoto, Japan) on the non-dominant arm after participants after five minutes of sitting. Height, weight, waist and hip circumferences, and blood pressure measurements were conducted twice and averaged for analysis. Body composition indices (fat free mass (FFM), fat mass, and their percentages) were assessed using bioelectrical impedance analysis (BIA, RJL-Systems Quantum – V Segmental, Ref: Q5S) in a reclining position.

#### 2.4.4. Dietary Outcomes

Dietary data were collected using the NCI’s semi-quantitative past month version of the Diet History Questionnaire III (DHQ III) before and after the 6-week intervention [32]. Total and detailed DHQ nutrient compositions were analyzed and used to calculate Healthy Eating Index (HEI), Dietary Approaches to Stop Hypertension (DASH), and MED dietary pattern indices with the dietaryindex package in R [33] to provide important contextual information regarding dietary patterns. Self-administered single 24-hour dietary recalls using ASA24^®^ Dietary Assessment Tool [34] were also conducted before and after the intervention.

#### 2.4.5. Free-Living Appetite Ratings

The intensity of hunger, fullness, desire to eat, and prospective consumption were measured on electronic versions of 100-mm visual analog scales, with scale endpoints ranging from “not at all” to “extremely” [31]. Participants recorded their responses every hour during the waking hours in a 24-h period. The time points between 8 am to 8 pm had the most complete data and were considered for further analysis. Intermittent missing time points within an assessment day were imputed within participant using neighboring observed values (carried forward or backward or averaged when both were available) to enable calculation of within-day AUCs. Morning, afternoon, evening, and total 12-hour period area under the curve (AUC) indices were computed using the trapezoidal rule.

#### 2.4.6. Acceptance and Palatability Ratings

A 9-point food action rating scale assessed the weekly acceptability of almond and cookies (1 = “I would eat this if I were forced to” and 9 = “I would eat this every opportunity I had”) [35]. Additionally, the palatability of food items was evaluated weekly by a hedonic general labelled magnitude scale (gLMS) (100 = “Extremely unpalatable” and 100 = “Extremely palatable”) [36].

### 2.5. Statistical Analyses

Statistical analyses were conducted in R (version 4.5.2). Linear mixed model analyses were performed using the lmerTest package [37] with group (Almond and Cookie) as the between-subject fixed effect, week (Baseline and Week 6) as the within-subject fixed effect, the group x week interaction, and participant included as a random intercept to account for repeated measures. For serum markers assessed via ELISA, the plate number was included as a covariate to control for variations according to plates. For dietary outcomes, energy intake was included as a covariate. Anova type III tests were conducted on the fitted model using the car package. When a significant week x group interaction was observed, estimated marginal means and planned contrasts were calculated using the emmeans package [38] to perform the following pairwise comparisons: (1) almond vs. cookie at baseline, (2) almond vs. cookie at week 6, (3) baseline vs. week 6 within the almond group, (4) baseline vs. week 6 within the cookie group, and (5) change from baseline to week 6 in the almond group vs. change from baseline to week 6 in the cookie group. P-values were adjusted for multiple comparisons using the multivariate t distribution (mvt) adjustment [39]. Baseline adjusted linear regression models were also fit using the lm() function in R to evaluate group differences at Week 6 while adjusting for baseline values of the outcome.

Missing outcome data were not imputed for linear mixed models, as maximum likelihood estimation inherently accommodates incomplete repeated-measures data under the missing-at-random assumption [40]. For baseline adjusted analyses, missing values were addressed using multiple imputation by chained equations implemented with the mice package using predictive mean matching [41]. Imputation was performed separately within each treatment group and included sex, race, age, baseline BMI, and the respective baseline value of each outcome as predictors as has been done by others [42]. The number of imputations was determined using the von Hippel rule based on the maximum observed proportion of missing data [43]. Imputed datasets were generated with 50 iterations per chain, analyses were conducted within each imputed dataset, and fixed-effect estimates were pooled using Rubin’s rules [44]. Estimated marginal means, contrasts, and joint tests were subsequently computed from the pooled model estimates

Model assumptions were evaluated using residual diagnostics, including skewness and kurtosis, Levene’s tests, and graphical inspection of residual distributions. Data not meeting normality assumptions were transformed using the Johnson’s family of transformations [45] in JMP Pro (version 17.0), however, only the non-transformed data are presented for interpretation purposes unless otherwise indicated.

The group effect on total compliance % was analyzed by standard least squares regression. A GEE-based multinomial logistic regression was conducted in R using the multgee package [46] to evaluate whether the timing of consumption (morning, afternoon, evening) differed by group, accounting for repeated days of data per participant. FACT and palatability data were analyzed using nonparametric analysis of week (W1, W2, W3, W4, W5, and W6) and group interaction effects using the nparLD package in R [47]. For non-parametric analyses, post-hoc tests for significant effects were conducted using Mann Whitney analyses for between-subject factors, and nparLD for within-subject factors, with adjustments across contrasts (comparions) using Benjamini-Hochberg correction.

## 3 Results

### 3.1 Participants Anthropometric, Cardiovascular, and Biochemical Characteristics

There were no group differences in body weight, waist and hip circumferences, blood pressure, serum glucose, insulin, insulin sensitivity and resistance indices, tocopherol (alpha and beta-gamma) concentrations, and serum lipid profile (total cholesterol, LDL-C, and triglycerides) over the 6-week intervention (**Table 2**). Although there were statistically significant group x week interaction effects for total fat mass %, total fat-free mass %, none of the post-hoc comparisons were significant. HDL-C concentrations were higher in the almond group than in the cookie group at W6 (BL-adjusted group effect, P<0.05); however, this difference was largely driven by a tendency for HDL-C to decline in the cookie group (Group x Week Effect, P ≤ 0.1).

**Table 2.**
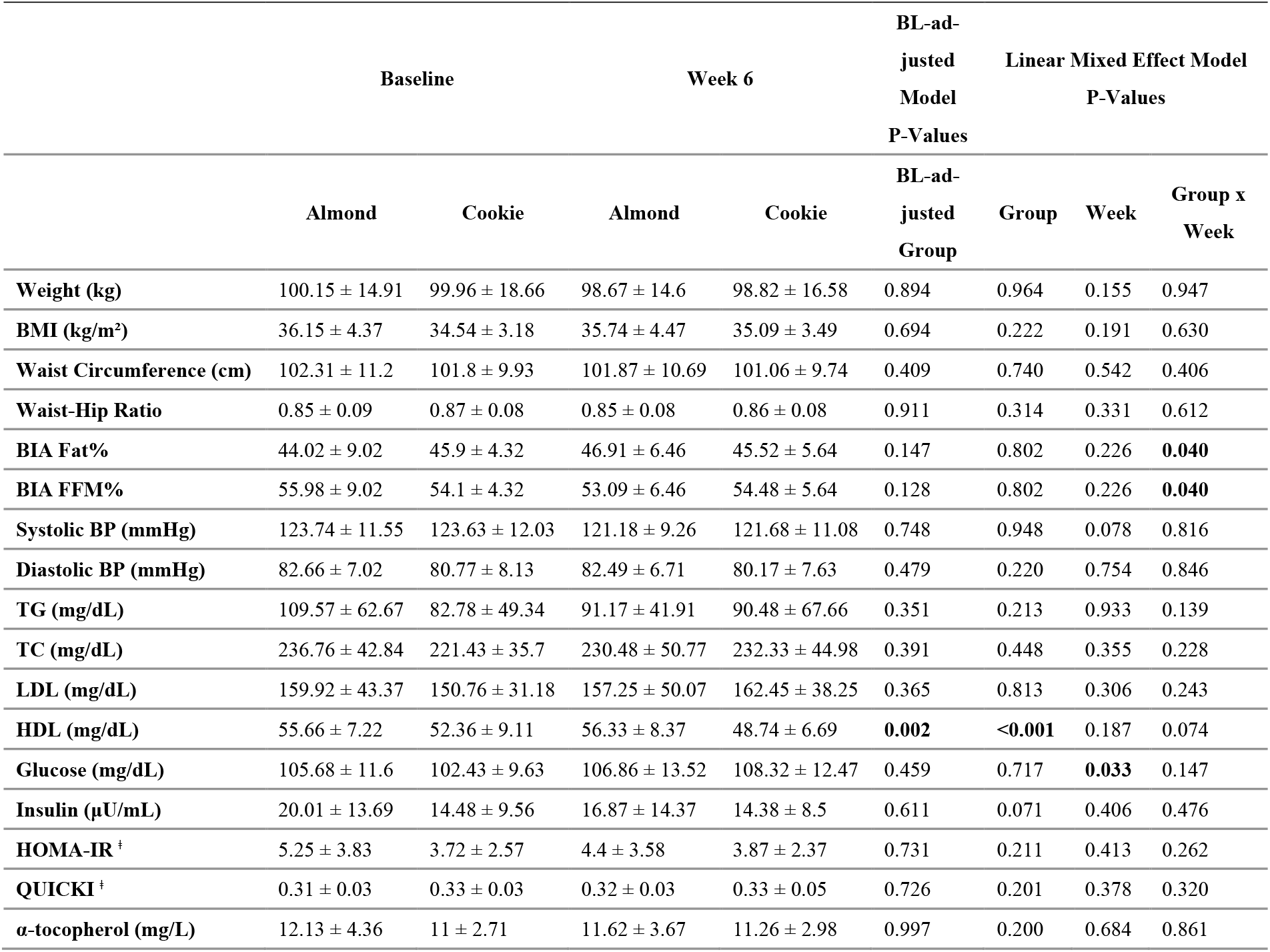
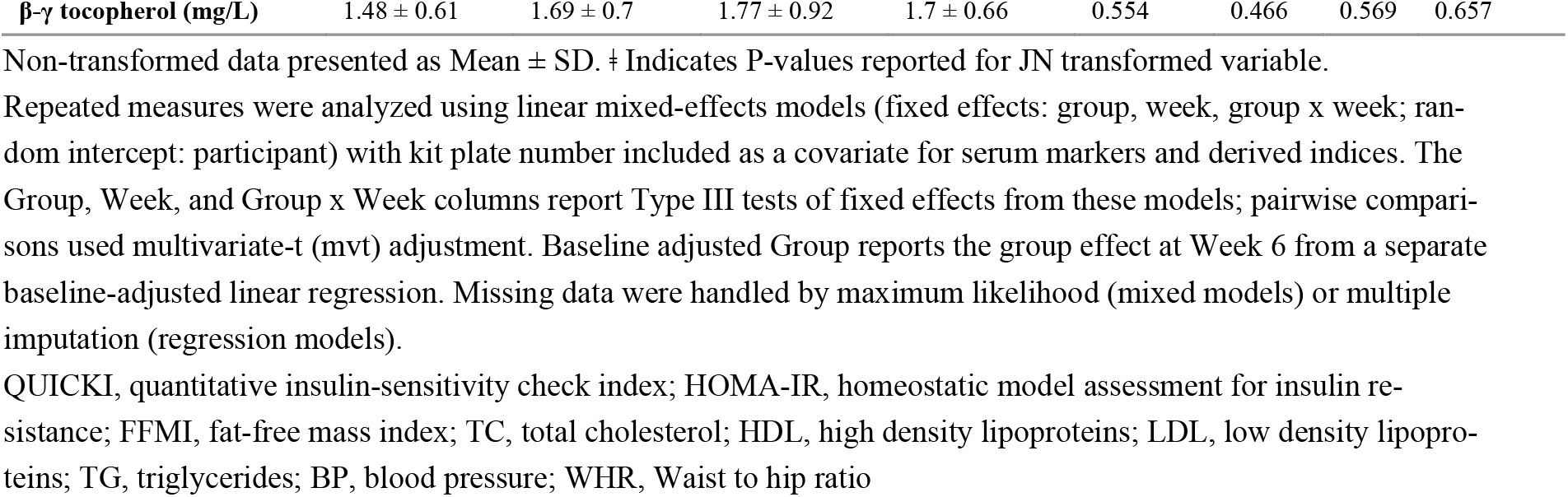
Anthropometric, body composition, and clinical data for almond and cookie groups at baseline and week 6 in adults with obesity.

### 3.2 Study Food Compliance and Acceptability

Mean subjective compliance percentage was high for both groups (Almond: 93.9% and Cookie: 92.5%) and no statistically significant group differences were observed. There were no group differences in time of consumption as well. Because ratings were first collected after one week of food consumption, baseline (pre-intervention) values are not considered. Overall, almonds had higher acceptance than cookies (group effect, P<0.05) but similar palatability. Acceptance and palatability decreased over the subsequent weeks of the intervention (time effect, P<0.05, **Table 3**), with no group x week interaction, indicating that changes over time were similar between groups.

**Table 3.**
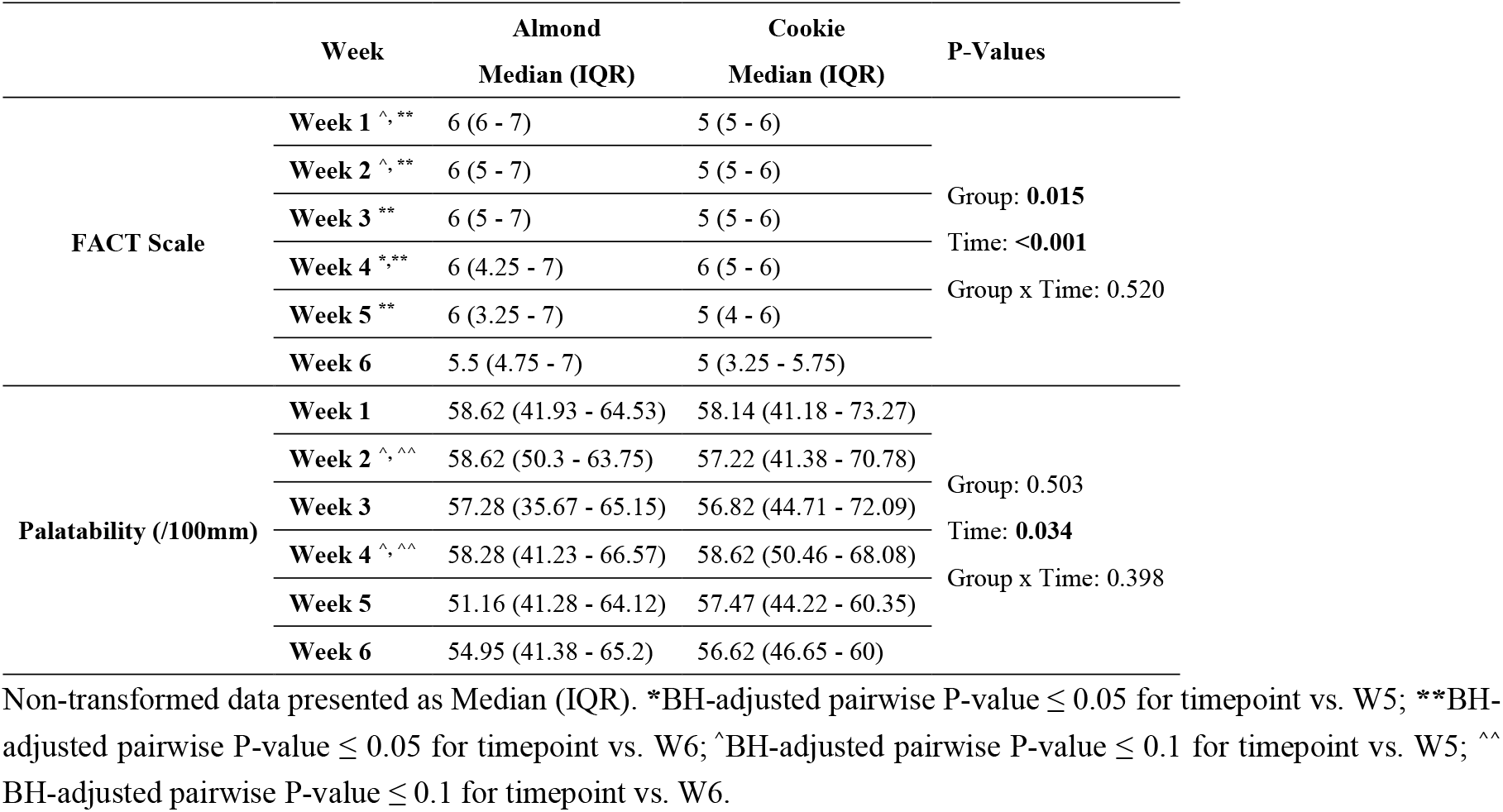
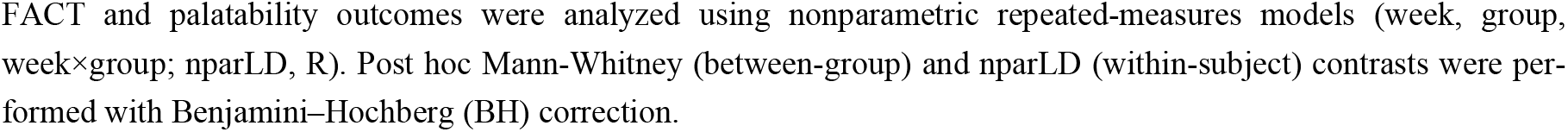
Acceptance and palatability ratings of study foods at baseline and week 6 in adults with obesity.

|  | Week | Almond<br>Median (IQR) | Cookie<br>Median (IQR) | P-Values |
| --- | --- | --- | --- | --- |
| FACT Scale | Week 1 <sup>^, **</sup> | 6 (6 - 7) | 5 (5 - 6) | Group: **0.015**<br>Time: **<0.001**<br>Group x Time: 0.520 |
|  | Week 2 <sup>^, **</sup> | 6 (5 - 7) | 5 (5 - 6) |  |
|  | Week 3 <sup>**</sup> | 6 (5 - 7) | 5 (5 - 6) |  |
|  | Week 4 <sup>*, **</sup> | 6 (4.25 - 7) | 6 (5 - 6) |  |
|  | Week 5 <sup>**</sup> | 6 (3.25 - 7) | 5 (4 - 6) |  |
|  | Week 6 | 5.5 (4.75 - 7) | 5 (3.25 - 5.75) |  |
| Palatability (/100mm) | Week 1 | 58.62 (41.93 - 64.53) | 58.14 (41.18 - 73.27) | Group: 0.503<br>Time: **0.034**<br>Group x Time: 0.398 |
|  | Week 2 <sup>^, ^^</sup> | 58.62 (50.3 - 63.75) | 57.22 (41.38 - 70.78) |  |
|  | Week 3 | 57.28 (35.67 - 65.15) | 56.82 (44.71 - 72.09) |  |
|  | Week 4 <sup>^, ^^</sup> | 58.28 (41.23 - 66.57) | 58.62 (50.46 - 68.08) |  |
|  | Week 5 | 51.16 (41.28 - 64.12) | 57.47 (44.22 - 60.35) |  |
|  | Week 6 | 54.95 (41.38 - 65.2) | 56.62 (46.65 - 60) |  |

FACT and palatability outcomes were analyzed using nonparametric repeated-measures models (week, group, week×group; nparLD, R). Post hoc Mann-Whitney (between-group) and nparLD (within-subject) contrasts were performed with Benjamini–Hochberg (BH) correction.

### 3.3 Appetite Ratings

No group differences in morning, afternoon, evening, or 12-hour AUCs were observed for hunger, desire to eat, or prospective consumption ratings over the 6-week intervention (**Table 4**). Although there were statistically significant group x week interaction effects for 12-hour fullness AUC, none of the post-hoc comparisons were significant.

**Table 4.**
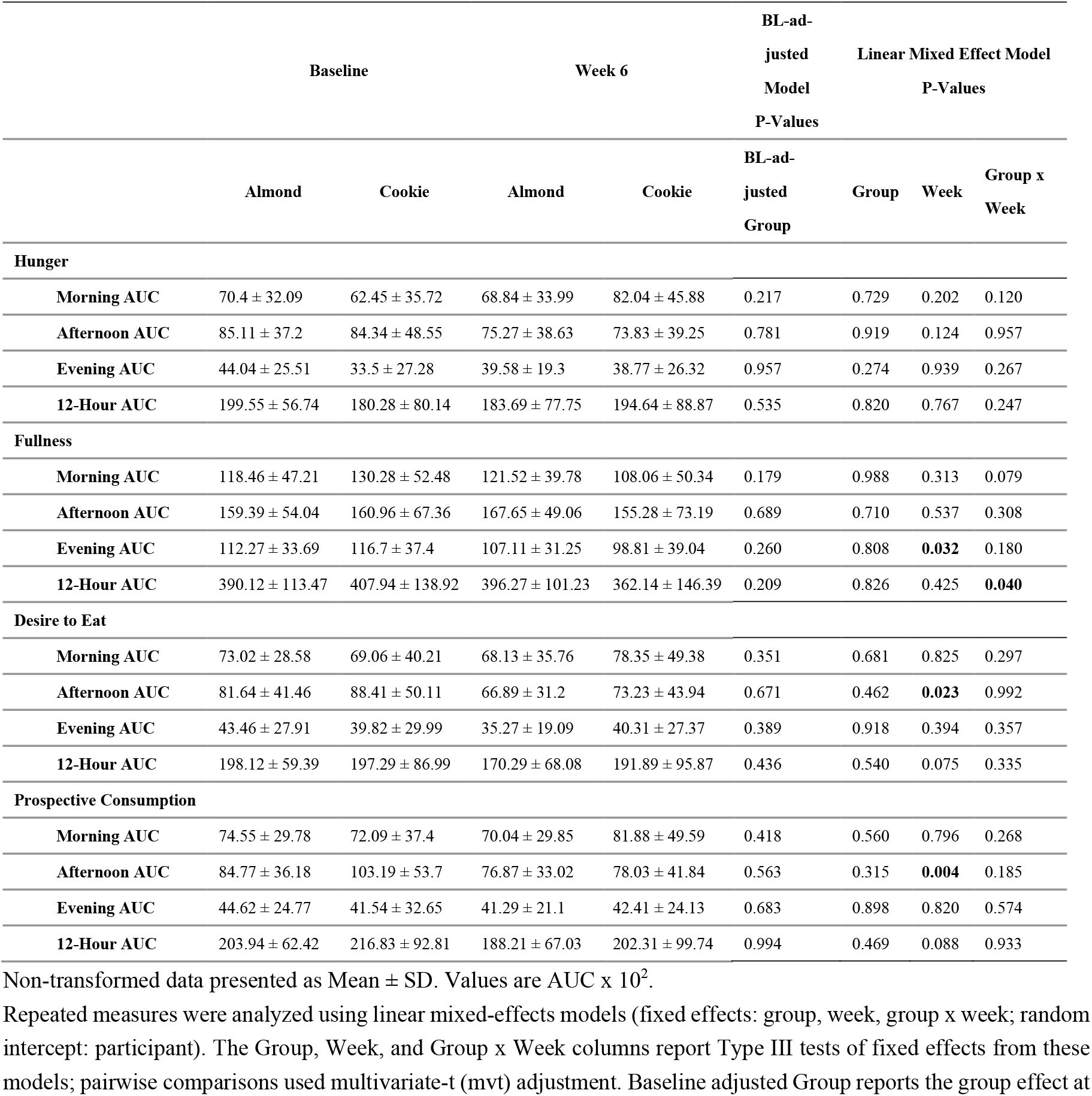
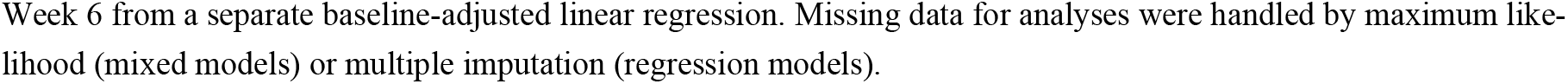
Subjective appetite ratings for almond and cookie groups at baseline and week 6 in adults with obesity.

|  | Baseline |  | Week 6 |  | BL-ad-justed Model P-Values | Linear Mixed Effect Model P-Values |  |  |
| --- | --- | --- | --- | --- | --- | --- | --- | --- |
|  | Almond | Cookie | Almond | Cookie | BL-ad-justed Group | Group | Week | Group x Week |
| **Hunger** |  |  |  |  |  |  |  |  |
| Morning AUC | 70.4 ± 32.09 | 62.45 ± 35.72 | 68.84 ± 33.99 | 82.04 ± 45.88 | 0.217 | 0.729 | 0.202 | 0.120 |
| Afternoon AUC | 85.11 ± 37.2 | 84.34 ± 48.55 | 75.27 ± 38.63 | 73.83 ± 39.25 | 0.781 | 0.919 | 0.124 | 0.957 |
| Evening AUC | 44.04 ± 25.51 | 33.5 ± 27.28 | 39.58 ± 19.3 | 38.77 ± 26.32 | 0.957 | 0.274 | 0.939 | 0.267 |
| 12-Hour AUC | 199.55 ± 56.74 | 180.28 ± 80.14 | 183.69 ± 77.75 | 194.64 ± 88.87 | 0.535 | 0.820 | 0.767 | 0.247 |
| **Fullness** |  |  |  |  |  |  |  |  |
| Morning AUC | 118.46 ± 47.21 | 130.28 ± 52.48 | 121.52 ± 39.78 | 108.06 ± 50.34 | 0.179 | 0.988 | 0.313 | 0.079 |
| Afternoon AUC | 159.39 ± 54.04 | 160.96 ± 67.36 | 167.65 ± 49.06 | 155.28 ± 73.19 | 0.689 | 0.710 | 0.537 | 0.308 |
| Evening AUC | 112.27 ± 33.69 | 116.7 ± 37.4 | 107.11 ± 31.25 | 98.81 ± 39.04 | 0.260 | 0.808 | **0.032** | 0.180 |
| 12-Hour AUC | 390.12 ± 113.47 | 407.94 ± 138.92 | 396.27 ± 101.23 | 362.14 ± 146.39 | 0.209 | 0.826 | 0.425 | **0.040** |
| **Desire to Eat** |  |  |  |  |  |  |  |  |
| Morning AUC | 73.02 ± 28.58 | 69.06 ± 40.21 | 68.13 ± 35.76 | 78.35 ± 49.38 | 0.351 | 0.681 | 0.825 | 0.297 |
| Afternoon AUC | 81.64 ± 41.46 | 88.41 ± 50.11 | 66.89 ± 31.2 | 73.23 ± 43.94 | 0.671 | 0.462 | **0.023** | 0.992 |
| Evening AUC | 43.46 ± 27.91 | 39.82 ± 29.99 | 35.27 ± 19.09 | 40.31 ± 27.37 | 0.389 | 0.918 | 0.394 | 0.357 |
| 12-Hour AUC | 198.12 ± 59.39 | 197.29 ± 86.99 | 170.29 ± 68.08 | 191.89 ± 95.87 | 0.436 | 0.540 | 0.075 | 0.335 |
| **Prospective Consumption** |  |  |  |  |  |  |  |  |
| Morning AUC | 74.55 ± 29.78 | 72.09 ± 37.4 | 70.04 ± 29.85 | 81.88 ± 49.59 | 0.418 | 0.560 | 0.796 | 0.268 |
| Afternoon AUC | 84.77 ± 36.18 | 103.19 ± 53.7 | 76.87 ± 33.02 | 78.03 ± 41.84 | 0.563 | 0.315 | **0.004** | 0.185 |
| Evening AUC | 44.62 ± 24.77 | 41.54 ± 32.65 | 41.29 ± 21.1 | 42.41 ± 24.13 | 0.683 | 0.898 | 0.820 | 0.574 |
| 12-Hour AUC | 203.94 ± 62.42 | 216.83 ± 92.81 | 188.21 ± 67.03 | 202.31 ± 99.74 | 0.994 | 0.469 | 0.088 | 0.933 |

Non-transformed data presented as Mean ± SD. Values are AUC x 10<sup>2</sup>.
Week 6 from a separate baseline-adjusted linear regression. Missing data for analyses were handled by maximum likelihood (mixed models) or multiple imputation (regression models).

### 3.4. Dietary Assessments

All nutrients and indices were adjusted for energy intake in the analyses (**Table 5** and **Table S2**). The energy-adjusted analyses of the past month DHQ data shows that total fat, total MUFA, oleic acid, total fiber, insoluble fiber, alpha-tocopherol, beta-tocopherol, calcium, magnesium, manganese, phosphorous, zinc, and dietary pattern scores for HEI, MED, and DASH were higher in the almond group, and delta-tocopherol, gamma-tocopherol, and refined grains intake were lower in the almond group compared to the cookie group at the end of the 6-week intervention (BL and Energy-adjusted Group Effect, P<0.05, **Table 5**). Although there were no BL-adjusted group differences in protein and potassium intake at W6, statistically significant group x week interaction effects indicate a decrease in protein and potassium intake in the cookie group over the 6-week intervention. The findings from the single ASA24 dietary recall data reflecting intake over the day preceding the visits at BL and W6 also reflected higher intake of MUFA, oleic acid, alpha-tocopherol, and phosphorous in the almond group at W6 (BL and Energy-adjusted Group Effect, P<0.05, **Table S2**) and a tendency for a decline in potassium intake in the cookie group (Group x Week Effect, P<0.05, **Table S2**).

**Table 5.**
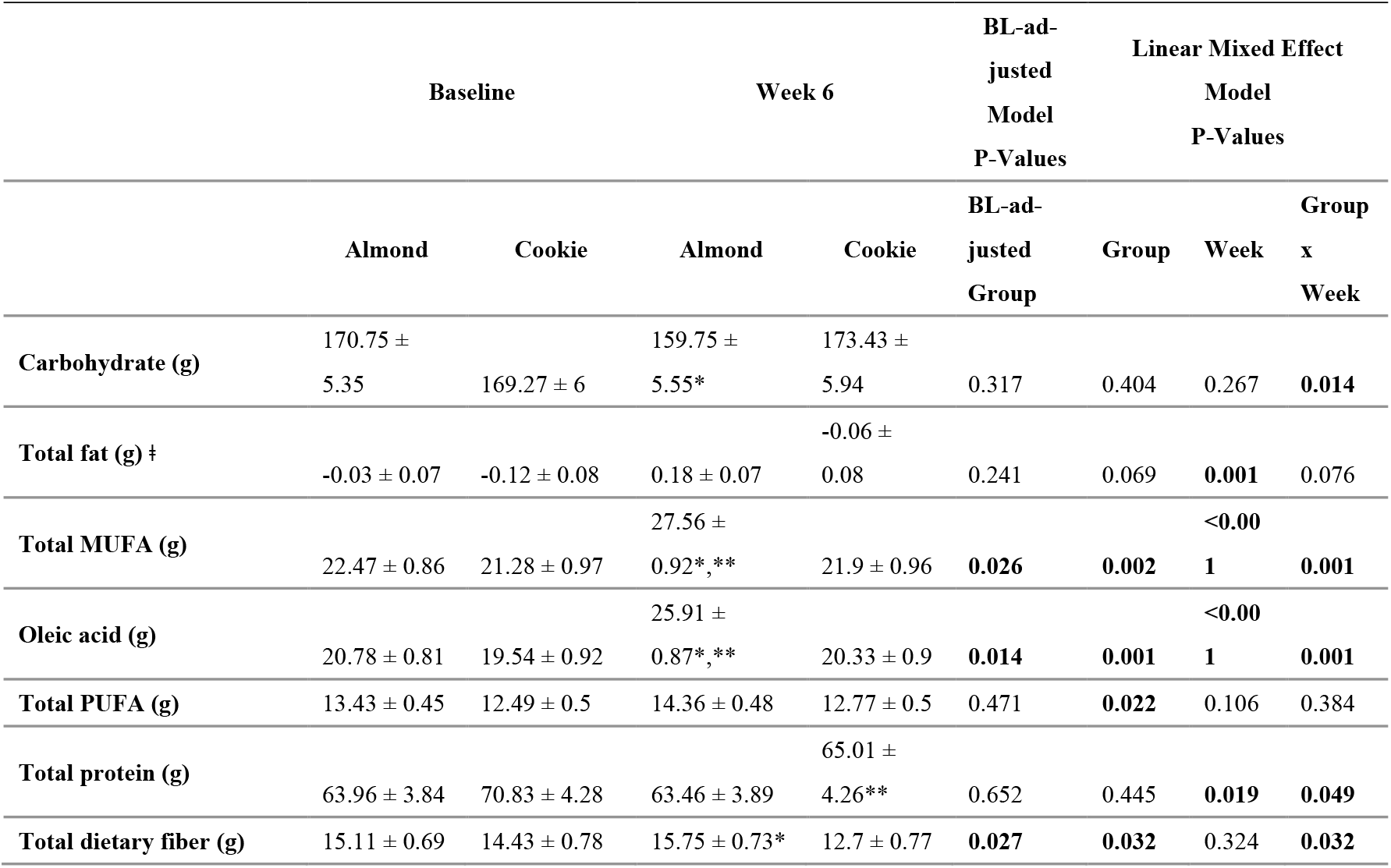
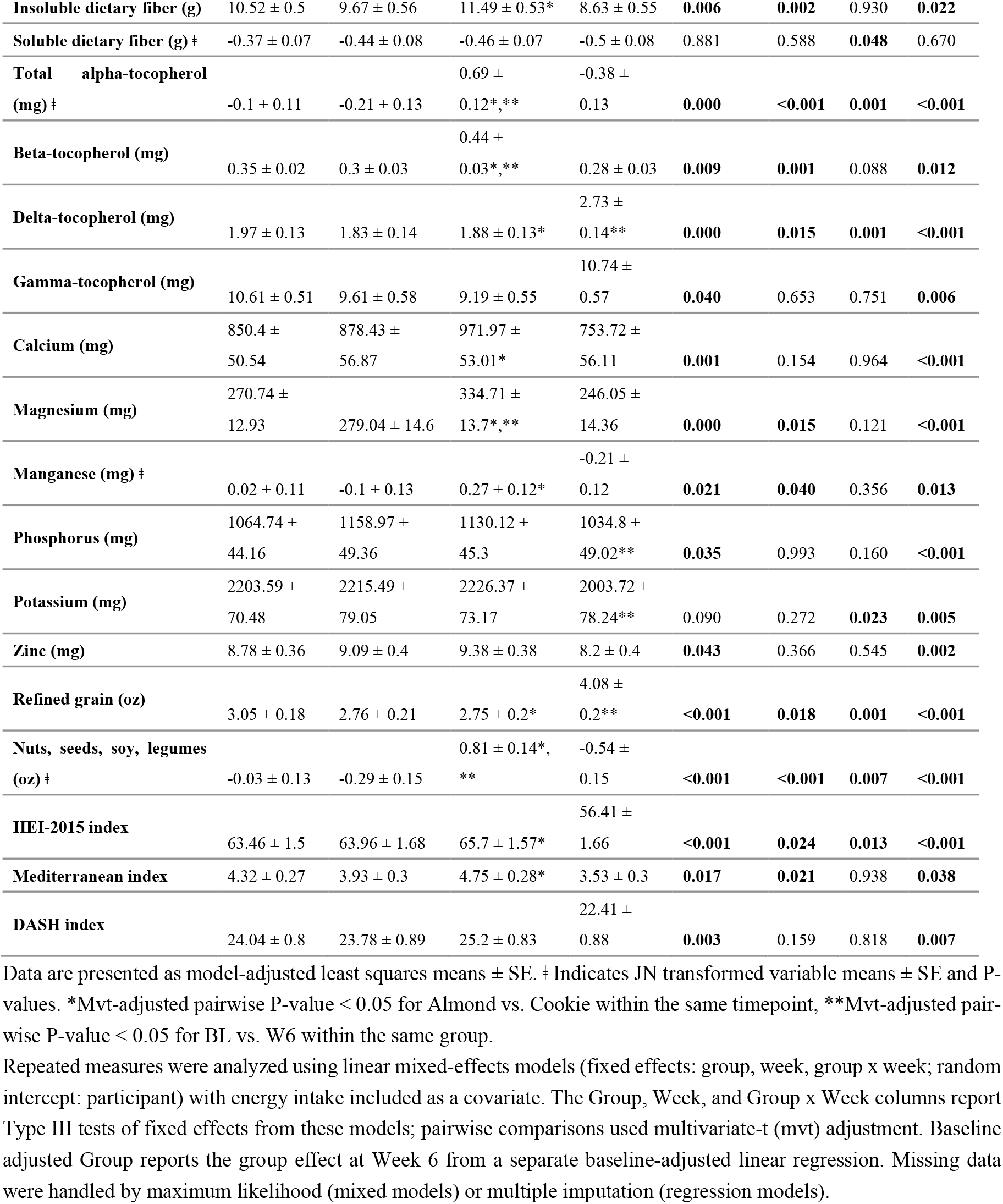
Self-reported dietary intake (energy-adjusted) from the Diet History Questionnaire III for the almond and cookie groups at baseline and week 6 in adults with obesity.

### 3.5 Immune and Inflammatory Marker Assessments

Overall, there were no group differences in immune markers over the 6-week intervention (**Figure 2**). However, almond consumption for 6 weeks resulted in lower concentrations of IL-6, TNF-alpha and IFN-gamma compared with cookie (BL-adjusted Group Effect, P<0.05 **Figure 3, Table S3**). No baseline-adjusted group differences were observed for the anti-inflammatory marker IL-10 at week 6; however, a statistically significant group x week interaction indicated that the increase in IL-10 over time was greater in the almond group compared with the cookie group. Analyses for IL-10 were conducted in a subset of participants due to quality control issues affecting two assay plates that rendered those data unusable, and results should therefore be interpreted cautiously due to reduced statistical power (**Table S3)**.

**Figure 2:**
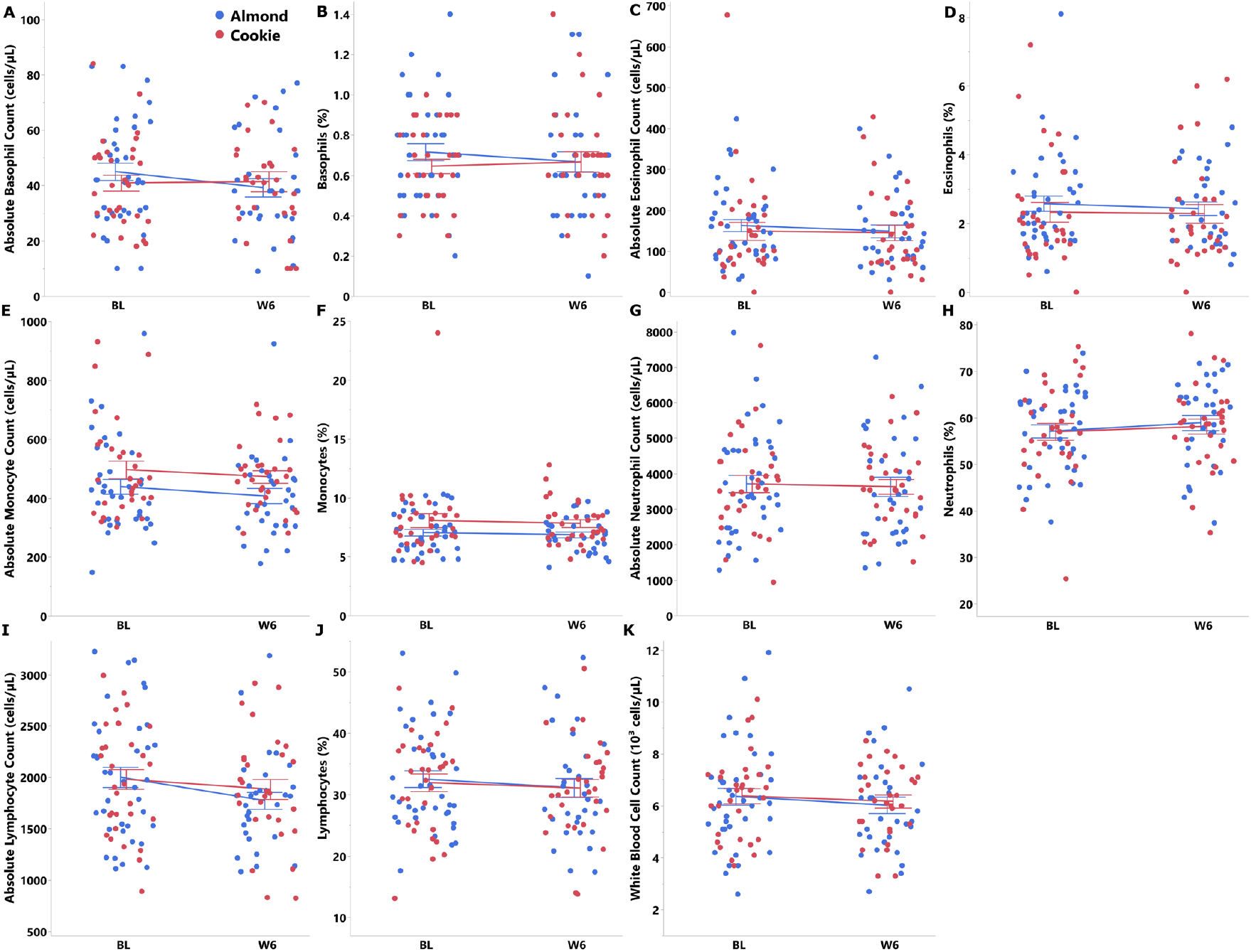
Immune markers in the almond and cookie groups at baseline and week 6 in adults with obesity. A: Absolute basophil count. B: Basophils (%). C: Absolute eosinophil count. D: Eosinophils (%). E: Absolute monocyte count. F: Monocytes (%). G: Absolute neutrophil count. H: Neutrophils (%). I: Absolute lymphocyte count. J: Lymphocytes (%). K: White blood cell count. Non-transformed data are presented as Mean ± SE. Repeated measures were analyzed using linear mixed-effects models (fixed effects: group, week, group x week; random intercept: participant) with kit plate number included as a covariate. The Group, Week, and Group x Week columns report Type III tests of fixed effects from these models; pairwise comparisons used multivariate-t (mvt) adjustment. Baseline adjusted Group reports the group effect at Week 6 from a separate baseline-adjusted linear regression. Missing data were handled by maximum likelihood (mixed models) or multiple imputation (regression models).

**Figure 3:**
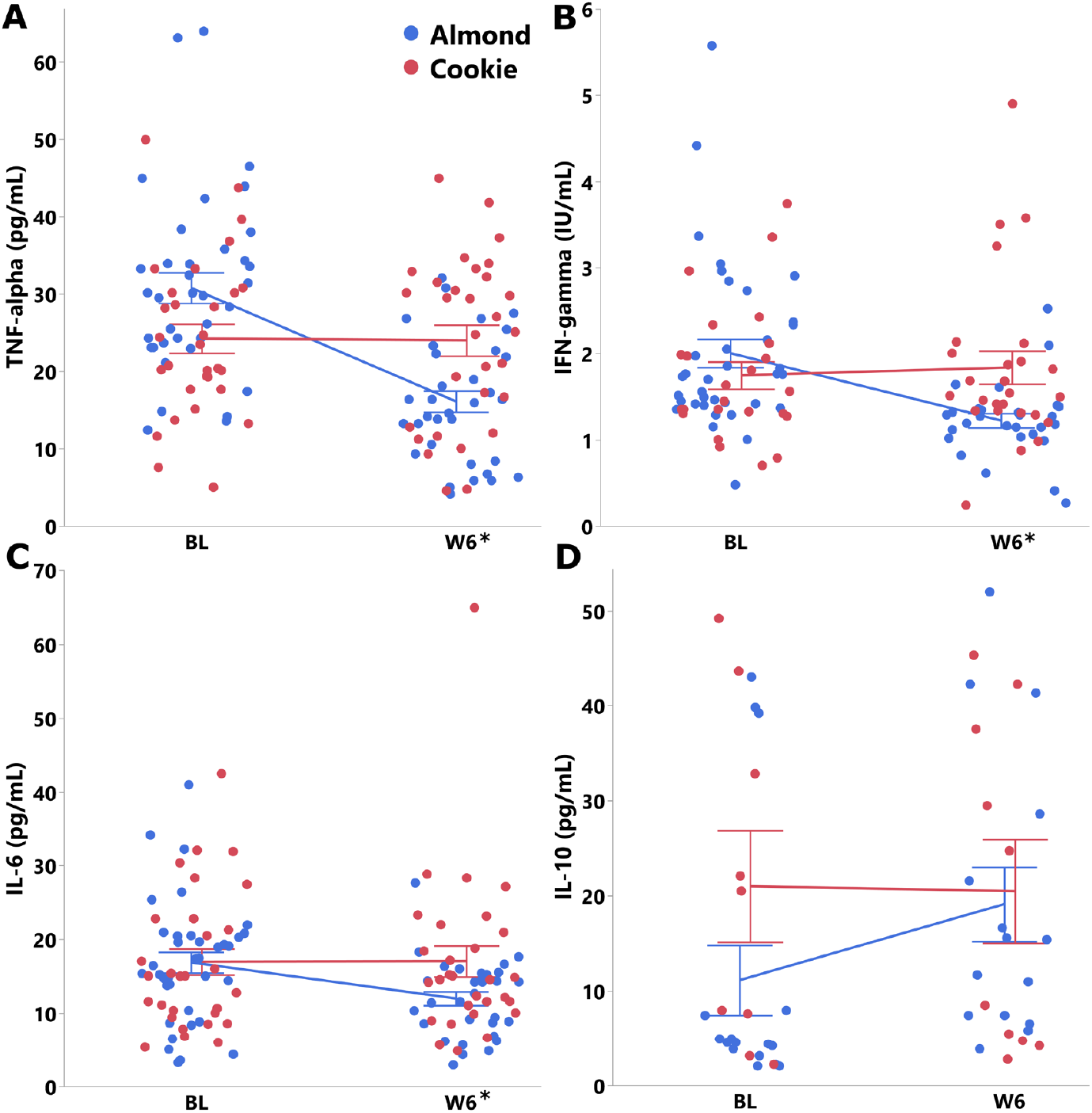
Inflammatory markers in the almond and cookie groups at baseline and week 6 in adults with obesity. A: TNF-alpha. B: IFN-gamma. C: IL-6. D: IL-10. Non-transformed data are presented as Mean ± SE. *BL-adjusted group effect P-value < 0.05. Repeated measures were analyzed using linear mixed-effects models (fixed effects: group, week, group x week; random intercept: participant) with kit plate number included as a covariate. The Group, Week, and Group x Week columns report Type III tests of fixed effects from these models; pairwise comparisons used multivariate-t (mvt) adjustment. Baseline adjusted Group reports the group effect at Week 6 from a separate baseline-adjusted linear regression. Missing data were handled by maximum likelihood (mixed models) or multiple imputation (regression models).

## 4. Discussion

Almond consumption for six weeks lowered serum concentrations of pro-inflammatory cytokines, including IL-6, TNF-α, and IFN-γ compared to an isocaloric ultra-processed snack of cookies. These improvements in inflammatory markers occurred under weight-maintenance conditions and were accompanied by improvements in overall diet quality and higher acceptance of almonds relative to cookies. Interestingly, other than the tendency of a decline in HDL-C with cookie consumption, this ultra-processed food did not impact other physiological or inflammatory markers. Since no changes in other cardiometabolic markers were observed over the six-week period, the pattern supports the interpretation that changes in low-grade inflammation may occur earlier and potentially precede detectable changes in cardiometabolic markers, which may require longer durations to manifest [48,49].

The observed reductions in IL-6, TNF-α, and IFN-γ, suggest that daily almond intake may help shift the balance of systemic inflammatory signaling toward a less pro-inflammatory state in adults with obesity. Previous meta-analyses examining the effects of almond consumption on inflammatory markers have yielded mixed results. One meta-analysis found that almond intake significantly reduced circulating CRP concentrations in adults across varying BMI and health statuses, but did not report obesity-specific subgroup effects [18]. Another found significant reductions in IL-6 overall, but these effects were more prominent in participants without obesity, however, with substantial heterogeneity observed within that subgroup [20]. A third meta-analysis found that while CRP and IL-6 were significantly lowered overall, these changes were not significant for individuals with obesity, and no significant effect overall was observed for TNF-α [19]. While the present study did not assess CRP, the magnitude and consistency of changes across cytokines (29–47% greater improvements in the almond group) suggest a stronger effect than previously reported in smaller or more heterogeneous populations. These effects were observed under free-living, weight-stable conditions, reinforcing that the nutrient composition of almonds, rather than energy restriction, mediated the cytokine shifts. These shifts are biologically plausible given both the specific nutrient changes and the overall improvements in diet quality documented in this study. Participants who consumed almonds had higher intakes of monounsaturated fatty acids, primarily oleic acid, which are known to reduce pro-inflammatory lipid mediators when they displace dietary saturated fats [50–52].

Almond consumption also increased intakes of key minerals, including magnesium, zinc, and manganese which synergistically may help support healthy immune and antioxidant responses that counteract chronic low-grade inflammation [58–60]. The almond group also had higher total fiber intake, driven mainly by the naturally high insoluble fiber content of almonds (approximately 80% of their total fiber (16)). Current evidence on how insoluble fiber influences inflammation, however, remains limited [61]. Collectively, these nutrient shifts were reflected in significantly higher HEI, MED, and DASH dietary pattern scores for the almond group compared to the cookie group. These dietary improvements with almond consumption likely acted through multiple complementary and synergistic pathways to produce the observed pattern of improved inflammation.

Notably, the almond intervention did not affect systemic innate or adaptive immune cell counts. This aligns with current understanding that obesity-related chronic inflammation is primarily driven by local immune cell infiltration and activation within adipose tissue which contributes to systemic inflammation [62–64], but does not necessarily cause large fluctuations in total circulating immune cell numbers over short periods. Although dietary patterns and weight loss can modulate this local immune environment and shift adipose tissue macrophage phenotypes [63], detecting corresponding changes in circulating blood counts may be challenging when there is no significant weight reduction, acute inflammatory challenge, or overt disease. While there are limited data on immune cells from nut-based studies, a systematic review found that vegetarian dietary patterns are associated with lower leukocyte counts in observational studies, yet intervention trials show inconsistent effects [65]. In one eight-week DASH diet study, reduced lymphocyte counts were observed, but these occurred alongside weight and fat loss, suggesting that adiposity change may mediate the effect [66]. Similarly, in a crossover trial, 4 week consumption of soy pudding modestly altered regulatory T cell counts, and in a 12-week RCT of a Brazilian cardioprotective diet with extra virgin olive oil, modest reductions in total leukocytes and lymphocyte-to-monocyte ratios were noted [67,68]. Collectively, these studies highlight that weight loss and intervention duration strongly influence total leukocyte counts, whereas shifts in immune cell subsets can occur with diet changes even without weight loss. Subtle immunological shifts may be better captured via adipose tissue analyses, gene expression profiling of isolated innate and adaptive cell subsets, and functional immune assays.

Almond consumption increased intake of alpha-tocopherol (vitamin E), a lipid-soluble antioxidant [53]. However, no significant differences in serum alpha-tocopherol concentrations were observed between groups over the six-week period. This is consistent with previous findings suggesting that modest dietary increases may not rapidly alter circulating levels in individuals with normal baseline concentrations due to tight homeostatic regulation and a saturable alpha-tocopherol pool [24]. Moreover, several factors may contribute to variability in circulating responses to intake, including substantial inter-individual heterogeneity in absorption efficiency (estimated ∼10–79%) [54], influences of baseline physiological and inflammatory status [55], and due to its fat-soluble nature, potential sequestration in liver fat [56], and greater partitioning into adipose tissue [57]. Collectively, these factors may attenuate detectable changes in serum concentration in individuals with obesity despite increased intake.

High compliance rates and higher acceptance scores for almonds compared to cookies underscore the feasibility of incorporating almonds into habitual eating patterns. Interestingly, cookies might have been expected to be more palatable due to their sweetness and ultra-processed characteristics, but there were no differences in palatability, and participants rated almonds as more acceptable overall. Appetite ratings remained stable with both almond and cookie consumption. Importantly, body weight remained stable in both groups, indicating effective energy compensation for both the ultra-processed cookie snack and almonds over 6 weeks. The findings are consistent with studies supporting that nuts do not contribute to positive energy balance when incorporated into a free-living diet [69–71].

The strengths of this study include its controlled design, high participant adherence, and stringent BMI and age criteria. However, the sample size, while adequate to detect changes in cytokine concentrations, may have limited power to detect changes in immune cell distributions. Other limitations pertained to quality control issues with the IL-10 assay, which precluded detection of true effects due to potential underpowering. The relatively short duration precludes conclusions about long-term sustainability of these effects. Finally, participants were generally healthy aside from obesity, and effects in populations with more advanced metabolic disease may differ.

In conclusion, this study provides new evidence that daily almond consumption can improve inflammatory profiles in middle-aged adults with obesity, independent of weight change and under free-living conditions. These results, together with the observed improvements in diet quality support including almonds as a practical component of dietary strategies to help attenuate obesity-related inflammation. Longer-term studies with larger samples may be needed to determine whether the observed improvements in inflammation mediate improvements in cardiometabolic outcomes in obesity. Additionally, functional immunological analyses and immunophenotyping could provide further insight into the mechanisms underlying almond-induced improvements in inflammation.

## Abbreviations

The following abbreviations are used in this manuscript:

DASH: Dietary approaches to stop hypertension
HEI: Healthy eating index
IFN: Interferon gamma
IL: Interleukin
MED: Mediterranean
TNF: Tumor necrosis factor

## Supplementary Materials

The following supporting information are available: Table S1: Nutrient composition of almond and cookie study snacks in the 6-week randomized controlled trial in adults with obesity; Table S2. Self-reported dietary intake (energy-adjusted) from the ASA-24 dietary recalls for the almond and cookie groups at baseline and week 6 in adults with obesity; Table S3. Immune and Inflammatory markers for the almond and cookie groups at baseline and week 6 in adults with obesity.

## Author Contributions

Conceptualization, J.D. and V.V.P.; methodology, J.D. and V.V.P.; investigation, J.D., P.B., and A.A.; formal analysis, J.D., A.A., and E.R.; data curation, J.D., A.A., and E.R.; writing—original draft preparation, J.D., A.A., and E.R.; writing—review and editing, all authors; supervision, J.D..; project administration, J.D.; funding acquisition, J.D. All authors have read and agreed to the published version of the manuscript.

## Institutional Review Board Statement

“The study was conducted in accordance with the Declaration of Helsinki and approved by the Institutional Review Board of University of Missouri-Columbia (protocol code 2090290 and date of approval: 05/11/2022).

## Informed Consent Statement

Informed consent was obtained from all subjects involved in the study.

## Data Availability Statement

Data described in the manuscript will be made available upon reasonable request pending completion of all study data analyses and publications.

## Acknowledgments

The computation for this work was performed on the high performance computing infrastructure operated by Research Support Solutions in the Division of IT at the University of Missouri, Columbia MO DOI: https://doi.org/10.32469/10355/97710

