## Supplementary Tables for "Almond Consumption Improves Inflammatory Profiles Independent of Weight Change: A 6-Week Randomized Controlled Trial in Adults with Obesity"

**Table S1. Nutrient composition of almond and cookie study snacks in the 6-week randomized controlled trial in adults with obesity**

|  | Almond^§^ | Cookie^¶^ |
| --- | --- | --- |
| **Weight, g** | 57 | 65 |
| **Energy, kcal** | 322* | 325 |
| **Carbohydrate, g** | 12 | 43 |
| **Dietary fiber, g** | 6 | 1 |
| **Protein, g** | 12 | 3 |
| **Total fat, g** | 30 | 16 |
| **Total SFA, g** | 2 | 5 |
| **Total MUFA, g** | 19 | 4 |
| **Total PUFA, g** | 7 | 6 |
| **Vitamin E (alpha-tocopherol), mg** | 11 | 1 |
| **Calcium, mg** | 120 | 14 |
| **Magnesium, mg** | 160 | 27 |
| **Iron, mg** | 2 | 4 |
| **Potassium, mg** | 434 | 113 |
| **Sodium, mg** | 1 | 205 |
| **Zinc, mg** | 2 | 0 |

^§^ Nutrient composition of natural, whole, unsalted, and dry roasted almonds provided by the Almond Board of California.

^¶^ Nutrient composition of Chips Ahoy chocolate chip cookies obtained from food label and the USDA food composition database.

*Metabolizable energy calculated as the average of Atwater general factor estimates and measured energy estimates from Gebauer et al. 2016.

**Table S2.** Self-reported dietary intake (energy-adjusted) from the ASA-24 dietary recalls for the almond and cookie groups at baseline and week 6 in adults with obesity.

|  | **Baseline** | | **Week 6** | | | **BL-adjusted Model  P-Values** | **Linear Mixed Effect Model  P-Values** | | |
| --- | --- | --- | --- | --- | --- | --- | --- | --- | --- |
|  | **Almond** | **Cookie** | **Almond** | **Cookie** | **BL-adjusted Group** | | **Group** | **Week** | **Group x Week** |
| **Carbohydrate (g)** | 239.85 ± 9.34 | 235.59 ± 9.98 | 202.87 ± 9.51** | 233.82 ± 10.31 | | 0.086 | 0.257 | **0.005** | **0.009** |
| **Total Fat (g)** | 92.29 ± 3.79 | 85.41 ± 4.05 | 103.55 ± 3.86 | 90.75 ± 4.18 | | 0.231 | **0.039** | **0.003** | 0.282 |
| **Total MUFA (g)** | 32.38 ± 1.48 | 28.56 ± 1.58 | 39.16 ± 1.51 *,** | 28.58 ± 1.64 | | **0.001** | **<0.001** | **0.004** | **0.004** |
| **Oleic acid (g)** | 30.51 ± 1.39 | 26.71 ± 1.48 | 37.07 ± 1.43 *,** | 26.9 ± 1.54 | | **0.003** | **<0.001** | **0.003** | **0.005** |
| **Total PUFA (g) ǂ** | -0.13 ± 0.12 | -0.12 ± 0.13 | 0.07 ± 0.13 | 0.2 ± 0.13 | | 0.613 | 0.618 | **0.024** | 0.591 |
| **Protein (g)** | 82.73 ± 5.59 | 95.63 ± 5.96 | 94.68 ± 5.71 | 85.57 ± 6.18 | | 0.061 | 0.783 | 0.832 | **0.012** |
| **Total fiber (g) ǂ** | 0.1 ± 0.12 | -0.01 ± 0.13 | 0.01 ± 0.13 | -0.1 ± 0.14 | | 0.761 | 0.439 | 0.417 | 0.991 |
| **Alpha-tocopherol (mg)** | 11.5 ± 1.19 | 9.23 ± 1.26 | 16.45 ± 1.22 | 10.29 ± 1.32 | | **0.004** | **0.002** | 0.005 | 0.067 |
| **Calcium (mg)** | 929.79 ± 78.81 | 985.31 ± 83.54 | 1044.67 ± 81.08 | 894.36 ± 87.15 | | 0.474 | 0.581 | 0.876 | 0.175 |
| **Magnesium (mg)** | 319.36 ± 21.19 | 329.25 ± 22.53 | 365.3 ± 21.69 | 303.3 ± 23.41 | | 0.084 | 0.302 | 0.577 | **0.041** |
| **Phosphorus (mg)** | 1357.08 ± 70.86 | 1479.21 ± 75.51 | 1540.84 ± 72.37 | 1311.89 ± 78.27 | | **0.040** | 0.538 | 0.884 | **0.002** |
| **Potassium (mg)** | 2500.77 ± 123.5 | 2628.93 ± 131.01 | 2551.35 ± 126.85 | 2224.42 ± 136.52 ^^ | | 0.176 | 0.477 | 0.121 | **0.043** |
| **Zinc (mg) ǂ** | -0.04 ± 0.11 | -0.07 ± 0.12 | 0.12 ± 0.12 | -0.02 ± 0.12 | | 0.571 | 0.429 | 0.373 | 0.620 |
| **Refined grains (oz. eq.)** | 5.36 ± 0.52 | 5.5 ± 0.55 | 5.53 ± 0.54 | 6.11 ± 0.58 | | 0.495 | 0.528 | 0.447 | 0.659 |

Data are presented as model-adjusted least squares means ± SE. ǂ Indicates JN transformed variable means ± SE and P-values. *Mvt-adjusted pairwise P-value ≤ 0.05 for Almond vs. Cookie within the same timepoint, **Mvt-adjusted pairwise P-value ≤ 0.05 for BL vs. W6 within the same group, ^^Mvt-adjusted pairwise P-value ≤ 0.1 for BL vs. W6 within the same group

Repeated measures were analyzed using linear mixed-effects models (fixed effects: group, week, group x week; random intercept: participant) with energy intake included as a covariate. The Group, Week, and Group x Week columns report Type III tests of fixed effects from these models; pairwise comparisons used multivariate-t (mvt) adjustment. Baseline adjusted Group reports the group effect at Week 6 from a separate baseline-adjusted linear regression. Missing data were handled by maximum likelihood (mixed models) or multiple imputation (regression models).

**Table S3.** Immune and Inflammatory markers for the almond and cookie groups at baseline and week 6 in adults with obesity.

|  | **Baseline** | | **Week 6** | | **BL-adjusted Model  P-Values** | **Linear Mixed Effect Model  P-Values** | | |
| --- | --- | --- | --- | --- | --- | --- | --- | --- |
|  | **Almond** | **Cookie** | **Almond** | **Cookie** | **BL-adjusted Group** | **Group** | **Week** | **Group x Week** |
| **Absolute basophils count (cells/uL)** | 45 ± 3.01 | 40.9 ± 3.33 | 41.6 ± 3.17 | 41.16 ± 3.36 | 0.384 | 0.584 | 0.362 | 0.289 |
| **Basophils %** | 0.72 ± 0.04 | 0.65 ± 0.05 | 0.67 ± 0.04 | 0.66 ± 0.05 | 0.554 | 0.480 | 0.660 | 0.295 |
| **Absolute eosinophils count(cells/uL)** | 162.68 ± 16.34 | 149.03 ± 18.09 | 157.79 ± 16.87 | 142.72 ± 18.18 | 0.808 | 0.535 | 0.430 | 0.921 |
| **Eosinophils %** | 2.58 ± 0.22 | 2.33 ± 0.25 | 2.42 ± 0.24 | 2.25 ± 0.25 | 0.976 | 0.497 | 0.351 | 0.750 |
| **Absolute monocytes count (cells/uL)** | 439.24 ± 23.87 | 496.84 ± 26.43 | 427.35 ± 24.83 | 472.73 ± 26.59 | 0.631 | 0.124 | 0.122 | 0.600 |
| **Monocytes %** | 7.06 ± 0.35 | 8.08 ± 0.39 | 6.83 ± 0.38 | 7.83 ± 0.4 | **0.038** | **0.029** | 0.354 | 0.954 |
| **Absolute neutrophils count (cells/uL)** | 3711.45 ± 228.95 | 3713.29 ± 253.49 | 3813.15 ± 239.31 | 3624.96 ± 255.21 | 0.549 | 0.770 | 0.955 | 0.426 |
| **Neutrophils %** | 57.09 ± 1.48 | 57.01 ± 1.64 | 58.47 ± 1.57 | 58.13 ± 1.66 | 0.729 | 0.919 | 0.151 | 0.885 |
| **Absolute lymphocytes count (cells/uL)** | 2001.15 ± 90.75 | 1980.84 ± 100.47 | 1920.78 ± 95.05 | 1887.12 ± 101.19 | 0.802 | 0.830 | 0.074 | 0.891 |
| **Lymphocytes %** | 32.53 ± 1.33 | 31.93 ± 1.47 | 31.6 ± 1.41 | 31.11 ± 1.49 | 0.907 | 0.763 | 0.280 | 0.945 |
| **White blood cells count (cells/uL)** | 6.36 ± 0.29 | 6.38 ± 0.32 | 6.38 ± 0.3 | 6.17 ± 0.32 | 0.445 | 0.812 | 0.470 | 0.365 |
| **TNF-alpha (pg/mL)** | 31.2 ± 1.69* | 25.17 ± 1.79 | 16.92 ± 1.72*,** | 24.86 ± 1.8 | **<0.001** | 0.600 | **0.000** | **<0.001** |
| **IFN-gamma (IU/mL)** | 2.32 ± 0.15 | 2.13 ± 0.19 | 1.61 ± 0.18** | 2.09 ± 0.17 | **0.006** | 0.305 | **0.009** | **0.019** |
| **IL-6 (pg/mL) ǂ** | 0.19 ± 0.17 | 0.19 ± 0.18 | -0.41 ± 0.18** | 0.19 ± 0.18 | **0.020** | 0.080 | **0.038** | **0.035** |
| **IL-10 (pg/mL) ǂ** | -0.12 ± 0.16 | 0.49 ± 0.2 | 0.64 ± 0.16** | 0.39 ± 0.19 | 0.109 | 0.318 | **0.012** | **0.001** |

Data are presented as model-adjusted least squares means ± SE. ǂ Indicates JN transformed variable means ± SE and P-values. *Mvt-adjusted pairwise P-value ≤ 0.05 for Almond vs. Cookie within the same timepoint. **^^^**Mvt-adjusted pairwise P-value ≤ 0.1 for Almond vs. Cookie within the same timepoint. **Mvt-adjusted pairwise P-value ≤ 0.05 for BL vs. W6 within the same group.

Repeated measures were analyzed using linear mixed-effects models (fixed effects: group, week, group x week; random intercept: participant) with kit plate number included as a covariate for serum markers and derived indices. The Group, Week, and Group x Week columns report Type III tests of fixed effects from these models; pairwise comparisons used multivariate-t (mvt) adjustment. Baseline adjusted Group reports the group effect at Week 6 from a separate baseline-adjusted linear regression. Missing data were handled by maximum likelihood (mixed models) or multiple imputation (regression models).
